# Fear, Access, and the Real-Time Estimation of Etiological Parameters for Outbreaks of Novel Pathogens

**DOI:** 10.1101/2020.03.19.20038729

**Authors:** Nina H. Fefferman, Eric T. Lofgren, Nianpeng Li, Pieter Blue, David J. Weber, Abdul-Aziz Yakubu

## Abstract

Early analysis of outbreaks of novel pathogens to evaluate their likely public health impact depends on fitting predictive models to data gathered and updated in real-time. Both transmission rates and the critical *R*_0_ threshold (i.e. the pathogen’s ‘reproductive number’) are inferred by finding the values that provide the best model fit to reported case incidence. These models and inferred results are then the basic tools used for public health planning: how many people expected to be infected, at what scales of time and space, and whether potential intervention strategies impact disease transmission and spread. An underlying assumption, however, is that the ability to observe new cases is either constant, or at least constant relative to diagnostic test availability.

We present a demonstration, discussion, and mathematical analysis of how this assumption of predictable observability in disease incidence can drastically impact model accuracy. We also demonstrate how to tailor estimations of these parameters to a few examples of different types of shifting influences acting on detection, depending on the likely sensitivity of surveillance systems to errors from sources such as clinical testing rates and differences in healthcare-seeking behavior from the public over time. Finally, we discuss the implications of these corrections for both historical and current outbreaks.

## Introduction

Mathematical models of the progression of the spread of infectious disease provide the tools used for real-time decision making in public health planning and outbreak management. They allow us to predict the time course of spread within a population(Chowell et al. 2017; Perkins et al. 2016; van den Driessche and Watmough 2002), provide critical cost-benefit estimates (Dasbach et al. 2006; Hayman et al. 2017; Keeling et al. 2017; Purdy et al. 2004), and evaluate best practices for particular interventions (Andrews and Basu 2011; Andrews and Bauch 2016; Ferguson et al. 2003; Kretzschmar et al. 2004). When confronted with a novel, potentially virulent pathogen, there is a rush to parameterize models appropriately (Capaldi et al. 2012; Farah et al. 2014; Sebrango-RodrÍGuez et al. 2017; Tizzoni et al. 2012), getting real-time case incidence data from surveillance sources and fitting the models to it to determine the likeliest estimates for probabilities of transmission (i.e. infectiousness) and the basic reproductive number, *R*_0_, which provides a metric of epidemic potential (Anderson 1991; Chowell et al. 2006; Chowell et al. 2004). As a new outbreak unfolds, updated incidence data helps refine the parameter estimates, shifting our understanding of the nature of the threat in real time (Moore 2004; Sebrango-RodrÍGuez et al. 2017; Tizzoni et al. 2012). However, many of these models make an explicit assumption that detection of new disease incidence is a function of well-understood confounders that remain mostly invariant over the course of an outbreak, such as the probability of an infected person developing symptoms. There are known corrections for instances that violate this assumption of constant detectability, such as when clinical case definition criteria are revised (Green 1998; Santermans et al. 2016; Thursky et al. 2003) or when new more sensitive and/or specific diagnostic tests become available (Nouvellet et al. 2015; Villela 2017). These confounders, however, are features of the surveillance process itself, and may therefore be understood so long as there is sufficient incorporation of medical and public health practice in the interpretation of the models (Villela 2017). Critically, these surveillance-based step-function changes may not the be the only meaningful factors confounding our ability to accurately estimate incidence data over time, and therefore accurately model the progression of an outbreak.

The importance of incorporating human behaviors into predictive epidemiological models has gained attention over the past decade (e.g. (Bansal et al. 2007; Del Valle et al. 2005; Fenichel et al. 2011; Funk et al. 2010; Perra et al. 2011)). Many models have now explored the potential impact of behaviors that directly impact transmission (e.g. school closures (Earn et al. 2012; Ferguson et al. 2006; Gemmetto et al. 2014; Lofgren et al. 2008), social distancing (Glass et al. 2006; Maharaj and Kleczkowski 2012; Reluga 2010; Valdez et al. 2012), use of personal protective equipment (PPE) (Anderson and Garnett 2000; Duerr et al. 2007)), etc.). However, the impact of human behavior within the context of epidemic outbreaks is not limited only to those that affect the transmission patterns of the pathogen. Our functioning societies enter into an epidemiological observer effect (cf. (Dirac 1947)) in which various behaviors are likely to confound both the sensitivity and specificity of surveillance detection of disease incidence.

Media-fanned public apprehension can create an over-demand for clinical testing (Sharma et al. 2003), even in the absence of clinical signs or symptoms and when transmission from asymptomatic persons does not occur (Baxter 2010). Social stigmatization associated with illness can conversely cause many with symptoms to avoid healthcare providers, and hence diagnosis, for as long as possible in order to avoid social repercussions (e.g. as with HIV/AIDS patients (Chesney and Smith 1999; Kalichman and Simbayi 2003). Even with fully rational and cooperative behavior on the part of the general public, public health directives and media attention will affect physicians themselves, potentially drastically altering rates at which physicians order tests to provide clinical diagnosis rather than relying on palliative treatment without the need for diagnosis (Barras 2020; Cowie et al.). This effect has already been shown to scale disproportionately with the actual rate of incidence (though not the focus of the study, this can be inferred from Fig 2 in (Iowa 1998 Annual Report)).

Further compounding the potential for these behavioral effects to mislead our models, the behaviors themselves are likely to depend on perceived epidemic status of the population. Individuals may shift their behaviors as reported prevalence rises and falls out of fear, or lack thereof, whether warranted by epidemiological truths or not. Case fatality rates are calculated based both on reported deaths and estimated case incidence, potentially amplifying the feedback since death may be considered an even greater motivator to action than illness. This implies that, not only do we may need to correct our predictive models for the pattern of surveillance sensitivity over time, but also to have sensitivity itself depend on the current perceived prevalence of the disease. This may be even more critical in instances where estimated case incidence does not accurately reflect numbers of infections (i.e. when case fatality rates and infection fatality rates differ significantly). In effect, modeling efforts should be split into separate endeavors: one of curve fitting for observed incidence, and one of inferring from those curves the likely underlying, actual disease process.

To capture this coupled process of disease dynamics and disease detection, we consider a standard, simple epidemiological model, but incorporate the potential for errors derived from a variety of sources that confound our estimates of case incidence. We use these models to demonstrate how these corrections would alter our understanding of historical outbreaks, and then discuss some evidence that modern outbreaks are affected by the types of behavioral shifts that we consider.

## Methods/Model

We begin with a standard Susceptible-Infected-Recovered (SIR) system, however, we will examine both “real” process of actual pathogen spread (denoted by the subscript *a*), and a “perceived” or “measured” process (denoted by the subscript *m*). For simplicity sake, we will assume that correct diagnosis and treatment has no bearing on the duration of illness/recovery time, nor on the rates of transmission from infected to susceptible individuals. Although both of these are obviously false for most outbreaks, they allow us to highlight the processes and methods most relevant to our purpose here and are easily corrected in specific application to particular outbreaks in the future. We therefore assume that the recovery rate, *γ*, is the same in both the perceived and real processes (i.e. *γ*_*a*_ = *γ*_*m*_ = *γ*).

This therefore yields a “real” process of 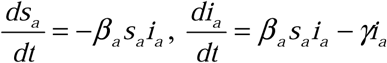, and *r*_*a*_ (*t*) *=* 1 − *s*_*a*_ (*t*) − *i*_*a*_ (*t*), where *s*_*a*_ (*t*), *i*_*a*_ (*t*), and *r*_*a*_ (*t*) are the fractions of the populations in the respective health categories at time *t*. To build the perceived disease process from this model, we then incorporate rates of testing for each fraction of the population, and the sensitivity and specificity of the test as follows.

Importantly, we will define as susceptible any person one who is not infected with our the pathogen of concern, despite possible infection with another illness. It is therefore not only reasonable but probable that “susceptible people” will seek out health care services and be tested for infection under our surveillance process, especially if the symptoms of their infection closely match those of the pathogen causing our focal outbreak. We therefore define *α* to be the rate at which susceptible people are tested for illness, call *δ* the rate at which infected people are tested for illness, and call *λ* the rate at which recovered people are tested for illness. (For purposes of this paper, we will assume *α = λ*, however this assumption may be relaxed in future work if memory of recently resolved symptoms affects health care seeking behavior). We define the false positive rate of the diagnostic test *ε* _1_ and the false negative rate of the test *ε* _2_(these may apply either to clinical diagnostic sensitivity and specificity, or else to error rates stemming from difference in physician opinion during syndromic surveillance).

Assuming that, at least initially, our surveillance cannot determine whether an uninfected person is susceptible or recovered, and therefore *s*_*m*_ (*t*) + *i*_*m*_ (*t*) *=* 1, we can define *s*_*m*_ = *s*_*a*_ (1 − *α*) + *s*_*a*_*α* (1 − *ε*_1_) + *i*_*a*_ (1 − *δ*) + *i*_*a*_*δ ε* _2_ + *r*_*a*_ (1 − *λ*) + *r*_*a*_ *λ* (1 − *ε*_1_) and *i*_*m*_ = *s*_*a*_ (*α ε*_1_ − *λ ε*_1_) + *i*_*a*_ (*δ* − *δ ε* _2_ − *λ ε*_1_) + *λ ε*_1_. Defined in this way, if *α = δ = λ=* 1, and *ε*_1_ = *ε*_2_ *=* 0, and *r*_*a*_ *=* 0, then *i*_*m*_ = *i*_*a*_ and *s*_*m*_ = *s*_*a*_ (i.e. when there are no errors and the surveillance is perfect, then the measured case incidence will be equal to the corresponding real case incidence, as we would hope).

Using this definition, we then correct our understanding of any disease incidence curve once we have either measured or assumed appropriate functions/values for *α, δ, λ, ε*_1_, and *ε*_2_. While this might at first seem straightforward, there arises the complication that our health care seeking behavior functions are likely to be problematic in at least three separate ways: (1) they are likely to be functions of the current perceived prevalence of infection in the population (i.e. some function of *i*_*m*_), (2) they are likely to be functions of time since the beginning of the perception of the current outbreak, (3) they are likely to be non-linear and, in some cases, not even continuous. We therefore propose the following algorithm to produce a system of SIR curves which reflect the underlying disease dynamics without the influence of behavioral shifts and/or testing inaccuracy; we will denote this system as “Testing Neutral”, TN.

We start from the most conservative assumption: that only the epidemiological rates of *β* _*m*_ and *γ* for the outbreak curve of interest are known (i.e. that the raw data to which an SIR model was fit to obtain those parameters is currently unavailable). We make this assumption to provide a method by which analysis of previously published rates for historical outbreaks could be analyzed without having to reanalyze the original outbreak data (should that data in fact be accessible, the correction can naturally be applied directly to the *i*_*m*_ data directly rather than to *i*\* curve described below). We, therefore, begin with an initially reconstructed SIR system (denoted by *) using only our measured *β*_*m*_ and 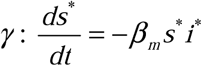 and 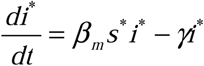. We then compute the corrected curve for the infected population (which is no longer necessarily continuous) using the definition of *i*_*m*_ above and applying it to the *i*\* and *i*_*cor*_ instead of *i*_*m*_ and *i*_*a*_ (respectively), we obtain 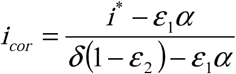 so long as 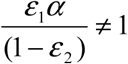 (note: if it is equal to 1, then *i*^*^ = *α*, which implies that the surveillance process cannot accurately capture the underlying real disease dynamics; derivation of this equality can be found in ESM Appendix 1). We are then able to generate the TN system by finding a new value of *β* which minimizes the square of the distance between the *i*_*cor*_ (*t*) curve and a new, hypothetical, standard continuous SIR system’s infected curve, using the known value of *γ*. We call this new, corrected value the “Testing Neutral *β* “which we denote *β*_*TN*_. So long as our assumed rates and behavior adjustment functions are reasonable approximations of the associated real-world values and behaviors, *β* _*TN*_ = *β* _*a*_, and the TN system may reasonably approximate the real disease dynamics (i.e. *s*_*TN*_= *s*_*a*_, *i*_*TN*_= *i*_*a*_, using the rates *β* _*TN*_and). These values of *β* _*a*_and (and by extension, the *R*_0_ computed either by fitting *i*_*TN*_ = *i*_*a*_, or else computed as the ratio of these corrected etiological rates) may then be compared to similarly corrected values for other outbreaks without worry that differences in sensitivity or health-care seeking behavior will influence the comparison.

## Results

### Demonstration of Impact of Healthcare-Seeking Behavior, Clinical Testing Rates, and Diagnostic Error Rates on Estimation of Outbreak Dynamics and Severity

To demonstrate the potential of these types of confounding factors in incidence estimation to influence our understanding of ongoing disease dynamics, we present the *i*_*m*_ and *i*_*a*_ curves under a variety of values for *ε*_1_, and *ε* _2_, and function choices for *α* and *δ*. Even under the simplest exploratory case, in which there are no ongoing dynamics affecting the ability to estimate incidence over time and where also the rates of testing for susceptible, infected, and recovered individuals are all held constant and identical, we see that asymmetry in error type rates alone can drastically alter our understanding of an ongoing outbreak (Fig. 1a). Extending this simple case to also include behavioral responses that shift over the course of an outbreak (i.e. non-constant testing rates), while still keeping all else the same, we see also that there can be drastic errors, even in the understood shape of the incidence curve to match the cases observed (Fig. 1b). (Again, for derivation of predictions for agreement/disagreement with real disease process based on the direction of the inequality between *A*_*ε*_ and 1, and the derivation of this example, see ESM Appendix 1).

**Figure 1:**
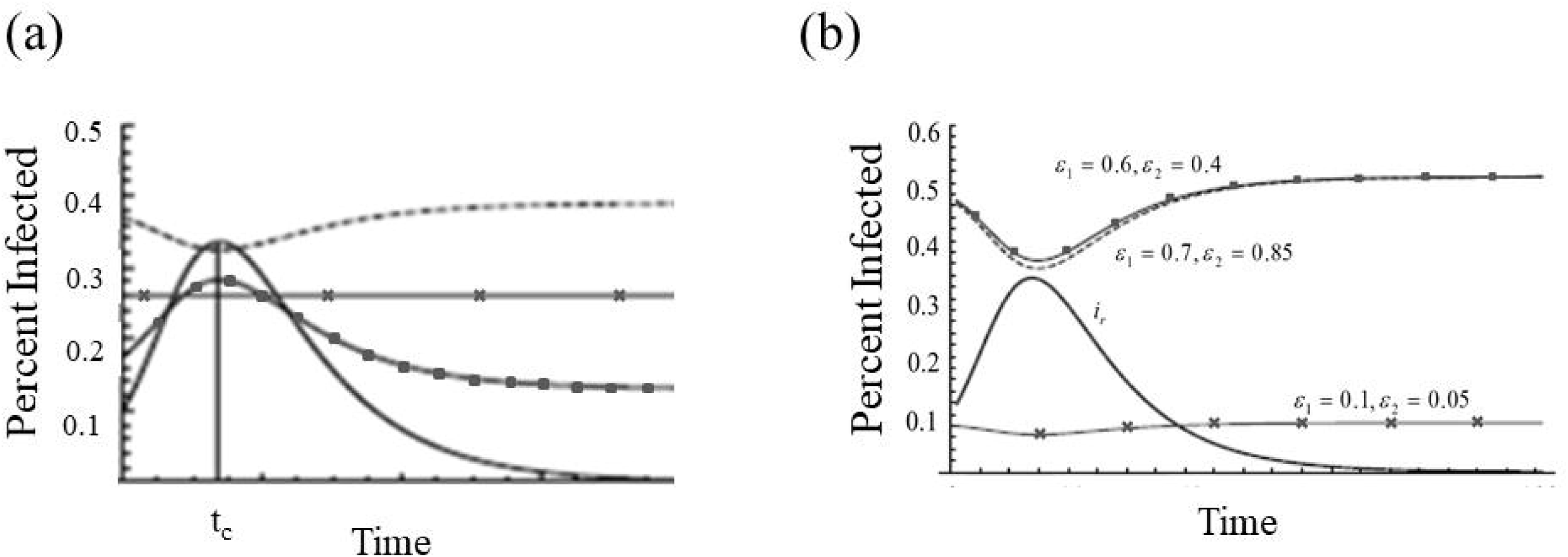
Example Perceived and Infected Curves Representing the Same Outbreak Under Different Testing Rates/Functions. All curves: *β* _*a*_ *=* 3, *γ* = 1, *s*_*a*_ (0) *=* 0.9, *i*_*a*_ (0) *=* 0.1. **(a)** Constant Behavioral Responses. Black solid curve: real disease dynamics; Black ■: *α* = 0.65, *δ* = 0.65, *ε*_1_ *=* 0.2, and *ε*_2_ *=* 0.1; Dashed curve: *α* = 0.65, *δ* = 0.65, *ε*_1_ *=* 0.6, and *ε*_2_ *=* 0.7 ; Black **×**: when the effective ratio of errors in testing, *A*_*ε*_ =1 (for calculations, see Appendix 1). **(b)** Non-Constant Behavioral Responses: All curves: *β* _*a*_ *=* 3, *γ* = 1, *s*_*a*_ (0) *=* 0.9, *i*_*a*_ (0) *=* 0.1. Black curve: real disease dynamics; All other curves *α* = 0.65, 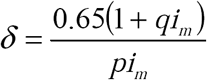, *p = q* = 1, (*ε*_1_ and *ε*_2_ for each curve as labeled).

Note that these calculations presented in Figure 1 are meant to be extremes to highlight the potential for confusion – we show a full range of values for *ε*_1_, and *ε* _2_ ranging from potentially realistic (*A*_*ε*_ =1) to dramatically inflated (both *ε*_1_, and *ε* _2_ are greater than 0.5, which would result in a more accurate test by simply negating the result). This is done to highlight the problem, though of course, real-world values are expected to be within a much narrower, more conservative range.

### Data-Driven Case Studies

#### Historical Outbreaks of Pandemic Influenza

Employing this now demonstrated potential for mismatch in understood dynamics to more realistic outbreak scenarios, we see that when health-care seeking behavior is dependent on the perceived prevalence of disease, shifting at a set threshold, there is also the potential for drastic misunderstanding of the disease dynamics, even if the error rates in testing are realistically low (Ai et al. 2020; Chu et al. 2012) (Fig 2a). Further departing from an idealized instructional case, when we incorporate both testing rate dependence on perceived prevalence and the amount of time since surpassing the threshold for increased behavioral demand for testing (e.g. gradual relaxation in public risk perception over time), the differences between the reality of the disease dynamics and the understanding that would be provided by fitting a model to case incidence data is even greater (Fig 2b).

**Figure 2:**
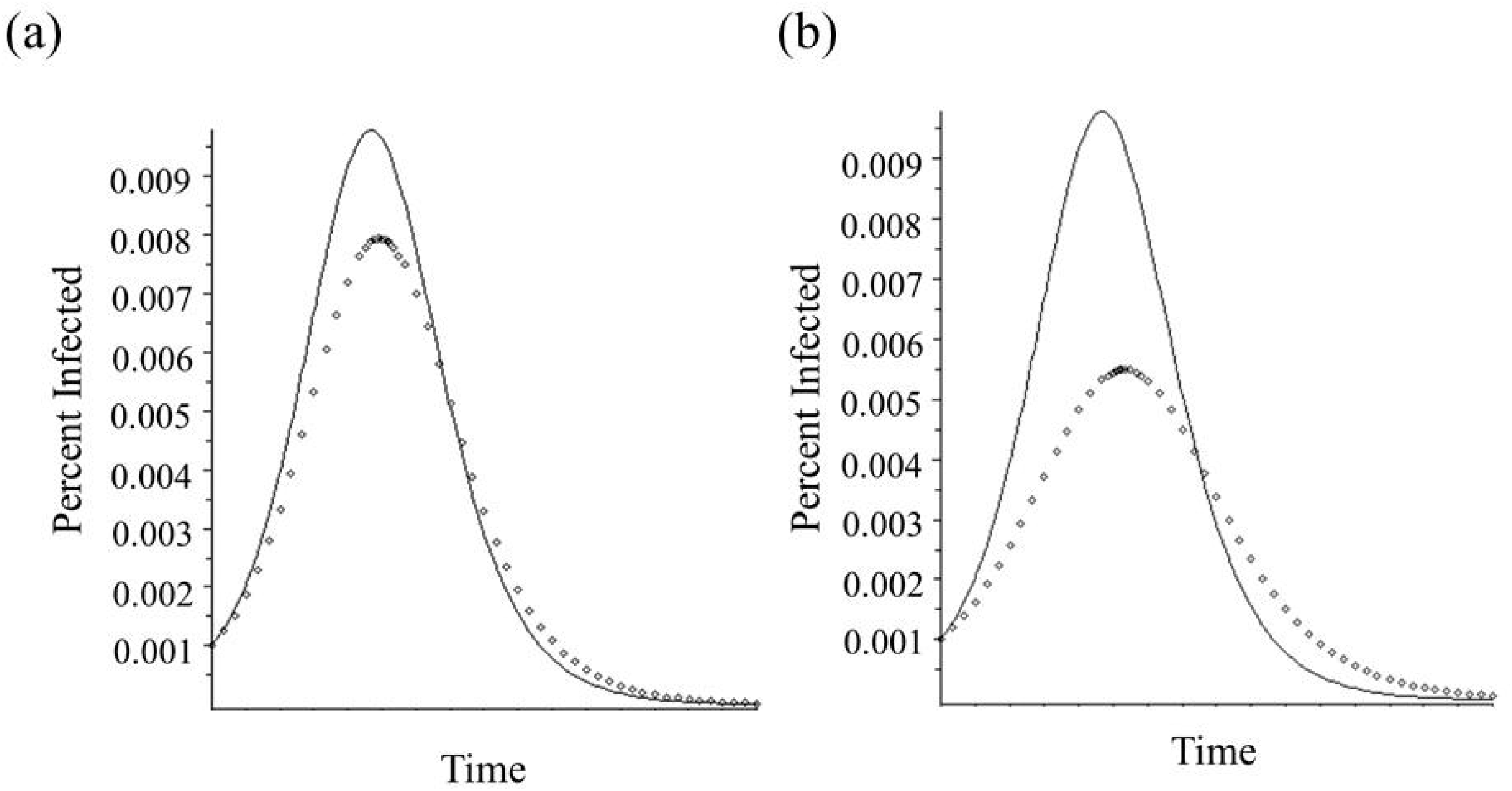
Example Perceived and TN Infected Curves Representing the Same Outbreak. **(a)** Non-Constant Health Care Seeking Behavior Functions. All curves: *γ* = 1, *s*_*a*_ (0) *=* 0.999, *i*_*a*_ (0) *=* 0.001. Solid curve – Perceived Outbreak: *β*_*m*_ =1.15, 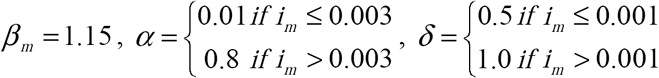, *ε*_1_ *=* 0.002, and *ε*_2_ *=* 0.005 ; Dotted curve – Testing Neutral Outbreak: *β*_*TN*_ *=* 1.13 **(b)** Healthcare Seeking Behavior Functions that Depend on Perceived Epidemic Severity and Time from first Outbreak Identification. All curves: *γ* = 1, *s*_*a*_ (0) *=* 0.999, *i*_*a*_ (0) *=* 0.001. Solid curve – Perceived Outbreak: *β*_*m*_ *=* 1.15, *α=* {0.01 if *i*_*m*_ has never exceeded 0.003, and 0.7 when *i*_*m*_ first exceeds 0.003, decreasing exponentially (by a factor of *e*^(*x* −*t*^)) over time to 0.3}, 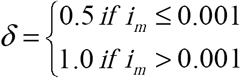, and *ε* _2_ *=* 0.005 ; Dotted curve – Testing Neutral Outbreak: *β*_*TN*_ *=* 1.10

To demonstrate how these effects might impact current understanding of modern analyses, we construct a hypothetical scenario using results from an excellent paper comparing the severity of pandemic and epidemic outbreaks of influenza: Viboud et al. 2006 (Viboud et al. 2006). In this paper, the authors concluded (among other things) that the *R*_0_ values for three pandemic years (1918, 1957, and 1968) were 2.1, 1.5 and 1.8 (respectively). However, while all three pandemic years of data were analyzed using transmission estimates inferred from influenza-attributed mortality data, the data for the 1957 and 1968 years were based upon WHO laboratory surveillance. For this reason, we can assume that the reported influenza attributed mortality was more accurate in representing only deaths from influenza (or associated pneumonia) than would have been possible for 1918. Entirely hypothetically, even if we assume that health care seeking behavior did not change at all between 1918 and 1957 (purely for demonstration, we assume 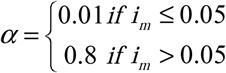 and 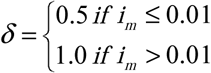 for all of these analyses), if we posit that the syndromic surveillance of 1918 led to error rates of *ε*_1_ *=* 0.1 and *ε*_2_ *=* 0.005, whereas the laboratory based testing was able to increase the specificity of the diagnosis (leaving the sensitivity the same) to *ε* _1_ *=* 0.01, we already see a drop in the perceived vs TN estimates of *R*_0_ for 1918 from 2.1 to 1.9, but no change (after rounding to the same number of digits) in the *R*_0_ estimates for either 1957 or 1968. This leads to a substantial mismatch in the observed incidence curve for the 1918 pandemic and an understanding of the same outbreak under a Testing Neutral assumption (Fig 3a) while both the 1957 and 1958 outbreaks would already have been accurately understood (Fig 3b and 3c).

**Figure 3:**
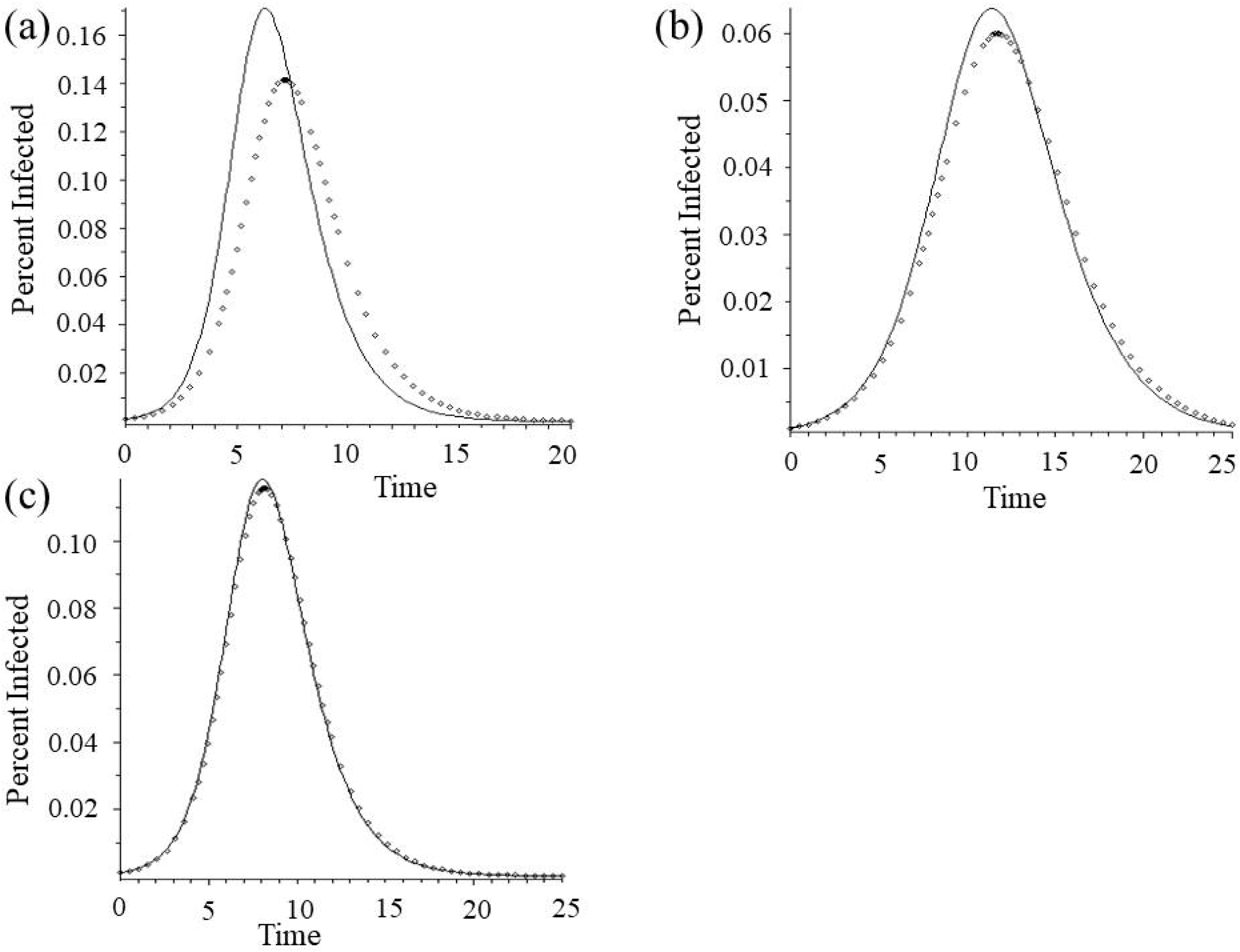
Differences in Estimates of *R*_0_ for Three Pandemic Years Using Hypothetical Correction Rates. **(a)** Analysis of Influenza Pandemic of 1918, **(b)** Analysis of Influenza Pandemic of 1957, **(c)** Analysis of Influenza Pandemic of 1968. For all panels – Solid line: Perceived/Reported pandemic incidence curve, reconstructed from reported *R*_0_. Dotted line: TN pandemic incidence curves (1918 TN *R*_0_ =1.9; TN 1957 *R*_0_ =1.5; 1968 TN *R*_0_ =1.8).

While we have no reason to suspect that our hypothetical error rates and assumed health care seeking behavioral functions reflect the reality of any of these three pandemics, they are clearly within realistic ranges and therefore demonstrate how dramatic the impact of even small differences in diagnostic sensitivity (whether due to changes in laboratory practice or to patient- or physician-driven behavior) can be on epidemiological estimates on which we base our public health strategies and policies.

#### Outbreak of Influenza H1N1-09

Whereas case studies of historical outbreaks of pandemic influenza allowed us to demonstrate the potential misestimate for *R*_0_ and resulting disease dynamics in the absence of direct understanding of behavioral shifts in testing practices, the more recent “novel” (H1N1-09) provides instead real-world data on the shifting demand for clinical diagnostic testing. This pandemic was first brought to light by global media attention *in advance* of clinical diagnosis in many areas. This is made clear by considering a time-series of both ordered clinical tests and confirmed cases of H1N1 in the UNC healthcare system in 2009 in which testing started immediately after media attention to the virus, but significantly before any actual circulation was detected (Fig. 4a).

**Figure 4:**
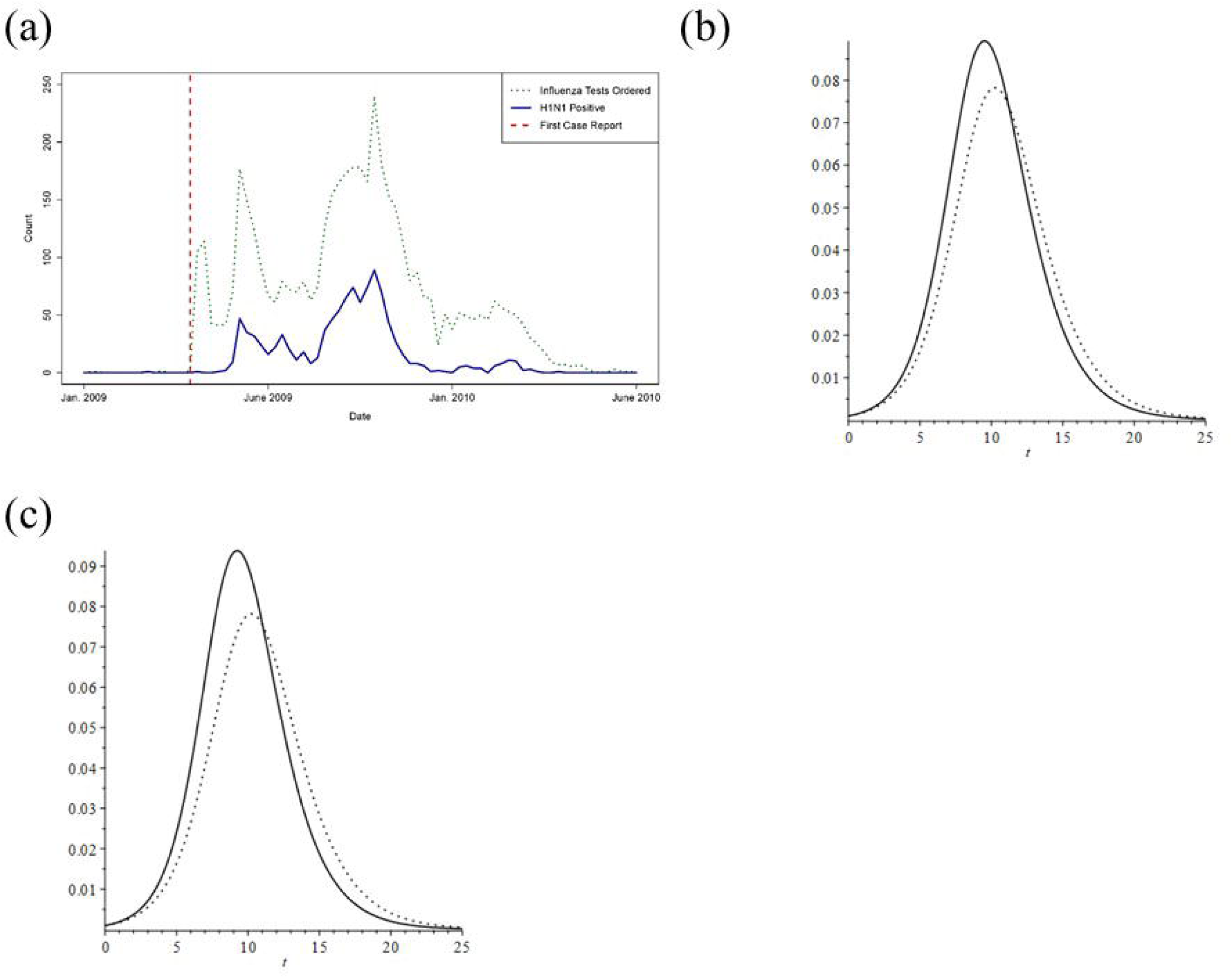
Testing Rates and Resulting Estimates of *R*_0_ for Novel H1N1 2009. **(a)** Counts of influenza tests ordered and H1N1 positive tests from UNC, **(b)** Estimated epidemic curves from reported (solid line) and TN (dotted line) epidemic incidence curves using the full time series, **(c)** Estimated epidemic curves from reported (solid line) and TN (dotted line) epidemic incidence using only the first 7 days of data after the first reported case to approximate real-time parameter estimation and resulting prediction.

Using the actual sensitivity and specificity known for the H1N1 tests in use at the time (Ginocchio et al. 2009), and the UNC testing curve to parameterize demand, we see that the reported estimate of *R*_0_ *=* 1.58 (Fraser et al. 2009), under correction, instead becomes and *R*_0_ *=* 1.64 (Fig. 4b). Of potential note, if we restrict the window for curve fitting to just the first weeks’ worth of data, we instead get an estimated *R*_0_ *=* 1.66 (Fig. 4c), meaning that, for this scenario, earlier estimates and projections were likely to overestimate the progression of the outbreak slightly. Depending on whether or not the UNC data is actually representative of broader patterns of test-seeking or test-ordering behavior this provides evidence that our understanding of the global dynamics of novel H1N1 in 2009 may be flawed.

#### Outbreak of COVID-19

While we have no way of currently estimating the rate of susceptible individuals seeking testing, we can make some generalizations given that the demand for testing in the United States as of 17 March well outstripped the supply of tests, and access to these tests was decidedly non-uniform (e.g. supplemental test availability from the Seattle Flu Study).

### Analytic Condition for Accuracy in Estimated Case Incidence from Surveillance

In addition to these numerical examples, we provide a theoretical threshold condition, *Ā*_*ε*_, for the ability of a surveillance system to reflect actual disease incidence based on assumed relationships among the behavioral functions and error rates (much as *R*_0_ provides a threshold condition for epidemics). Assuming that the behavioral health care seeking functions are independent of time, the effective ratio of error rates in the diagnostic tests, defined as 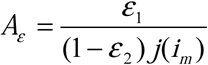, where *j*(*i*_*m*_) *=* ^*δ*^*/*_*α*_, can be used to define 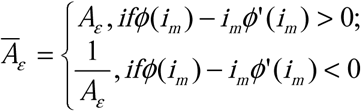 where *ϕ*(*i*_*m*_) = *α.* This *Ā*_*ε*_ then provides a way to determine whether the perceived or measured disease process may accurately reflect the real, underlying disease process. In this case, when the ratio of the diagnosis test rates is constant, if there are no errors in the diagnosis tests then the surveillance accurately reflects the real disease process though it may overestimate or underestimate actual incidence. If there are errors, the surveillance system accurately reflects the increasing or decreasing nature of the real disease if *Ā*_*ε*_ < 1, but can indicate increasing (resp. decreasing) incidence while the actual incidence is decreasing (resp. increasing) when *Ā*_*ε*_ > 1. When the ratio of the diagnosis tests is non-constant the results are more complicated, but some results are still accessible: without errors in diagnostic tests, a surveillance system can wrongly report no disease incidence while actual case incidence is either increasing or decreasing. Further, with small errors in the diagnostic tests it is possible for a surveillance system to report decreasing incidence while the actual incidence is increasing. (Proofs and characterizations of these relationships are provided in ESM Appendix 1.)

## Discussion

The ability to accurately infer epidemiological rates from outbreak data is critical to a majority of our public health planning efforts. As our models demonstrate, the accuracy of our estimates may be significantly compromised by our implicit assumption that diagnostic error rates and health care seeking behavior remain constant over the course of single, and even multiple, outbreaks, even as we know this assumption to be untrue. Regardless of the particular mechanism through which we attempt to characterize the changes in diagnostic sensitivity and specificity, our results demonstrate (in both theory and practice) how these dynamics may be incorporated into epidemiological modeling efforts and how the results may translate into a more accurate understanding of infectious disease dynamics.

Some studies have been able to assess the impact of public health announcement- or media-driven behavioral change with regard to disease risk and diagnosis (e.g. (Sharma et al. 2003)). It is clear that we will need to develop better models that explicitly capture the major factors that can effect change in public behavior regarding health care and diagnosis. While it may be impossible to accurately assess the impact of behavioral changes in health care seeking behavior for past epidemics, one possible course of action going forwards would be to ask physicians, hospitals and laboratories to record and report the number of tests performed in addition to merely the number of cases positively diagnosed, regardless of acknowledge threat of outbreaks.

These models and insights may also be of critical use our collective ongoing efforts to understand and predict the progression of COVID-19. Not only do we provide the obvious alternations to the standard epidemic predictions for error rates in testing, we also provide a mechanism by which to correct our understanding of *R*_0_ based on changes in access to tests of various sensitivities and specificities over time. This is especially important given both the formulation of governmental responses to the pandemic (i.e. “flattening the curve” or relying on community protection, *i*.*e*. ‘herd immunity’) and their subsequent evaluation hinge on accurate estimations of *R*_0_. While presented here with constant rates to enable the analytic calculations, real-time estimations of *R*_0_ are frequently based on numerical solutions, rather than analytic calculations. In this case, the expansion of precisely these equations to allow for *α, δ*, and *λ* to themselves be dynamic functions of public perception and disease prevalence will enable vastly more accurate understanding of real-time case incidence data. Work currently underway to try and capture the functional forms of these responses in observed behaviors in the US will hopefully allow us to extend these results very soon to the ongoing COVID-19 pandemic itself, but we provide this model in the meanwhile to allow others to work in parallel and improve our real-time decision-support capabilities.

## Data Availability

All data discussed in this manuscript is publicly available.

## Notes

### Competing Interest Statement

The authors have declared no competing interest.

### Funding Statement

This work was conducted without external funding support.

